# Temporal trends, socio-economic inequalities in obesity and responses by federal government, Nepal: a systematic review of observational studies, policies, strategies and plans, 2005-2019

**DOI:** 10.1101/2022.03.11.22272072

**Authors:** Ashok Bhurtyal, Dushala Adhikari

**Author notes:** Corresponding author Ashok Bhurtyal, Independent public health professional, Lalitpur, Nepal.

## Abstract

**Background:** Obesity has risen to epidemic proportions in low-income countries such as Nepal whose achievements in reducing maternal and child-undernourishment is well known. However, scientific evidence concerning recent transition towards obesity and corresponding responses by the state remains scanty. This review purported to assess the trends and disparities in obesity prevalence among women and children, and to analyse the governmental policies and programmes.

**Methods:** We searched PubMed and Google Scholar for articles published between January 1, 2005 and April 10, 2019, and websites of Demographic and Health Survey, Non-Communicable Diseases Risk Factor Survey, and Micronutrient Status Survey. We extracted data on the prevalence of obesity and overweight from the selected studies and synthesised narratively. Overweight and obesity prevalence data from the latest available nationwide surveys were disaggregated by gender, geographical location and household wealth quintile. We retrieved the federal governmental policies, strategies and plans from the websites of Ministry of Health and Population and the National Nutrition and Food Security Secretariat, National Planning Commission.

**Results:** Twenty studies that reported prevalence of overweight and/or obesity, with data from 79,082 men and women aged 15 years or more and 11,866 children under five years, were included. Obesity or overweight among men and women increased from 20.0% in 2004 to 36.1% in 2016 while obesity alone rose from 5.3% to 7.5%. Prevalence of childhood obesity or overweight remained very low, although doubled between 2006 and 2016, from 0.6% to 1.2%. Prevalences of overweight and obesity were much higher among women, inhabitants of urban areas and central provinces, and the affluent. Governmental policies, strategies and plans on nutrition were primarily designed to control undernutrition, with some direct and indirect implications for preventing obesity.

**Conclusions:** Prevalence of overweight and obesity increased substantially between 2005 and 2019, disproportionately affecting socio-economic and geographical groups in Nepal. Governmental efforts to contain the obesity epidemic should be reinforced by actions that are more specifically targeted to addressing obesogenic environments.

**Systematic review registration:** PROSPERO CRD42019132332

## Introduction

Obesity has become a major public health challenge worldwide, with rapid increments in prevalence of the malady in numerous low-income countries of the global south and south-east regions (1-5). Worldwide, prevalence of adult overweight and obesity, defined as a body mass index (BMI) of 25 kg/m^2^ more,, increased from 21-24% to nearly 40% between 1975 and 2016 (6). Although excess body weight was predominantly affecting wealthy countries in the past, the epidemic has deeply entrenched all parts of the world. However the occurrence of obesity differs substantially across countries and regions. Most of the low- or middle-income nations have witnessed a steady rise whereas several wealthier countries have observed a stabilisation or, at instances, reduction in the recent years (7-23). Obesity is associated with risks of chronic conditions such as cardiovascular diseases, type 2 diabetes, and some types of cancers (24, 25).

Although the decline in undernourishment is well established, obesity is a relatively lesser known phenomenon in Nepal. Over the past several years, the country has witnessed a steady reduction in maternal and child undernutrition attributed to a range of improvements in socio-economic and health sectors (26). However, this has come at the cost of increments in population obesity. Excess body weight accounts for 5% of mortality among Nepali people, which is much higher than proportion of deaths attributable to wasting (1.2%) or underweight (0.4%) among children (27). A number of studies have been conducted to assess the magnitude of overweight and obesity among Nepali people(28-30) including a review published a decade ago(31). Yet, a synthesised body of evidence concerning Nepal’s obesity epidemic remains unavailable.

We aimed to address this gap by assessing Nepal’s trends in prevalence of overweight and obesity among children and adults over the past one and half decades ; analysing disparities across gender, geographical locations and household wealth; and taking stock of governmental nutrition policies, strategies and plans for their significance in preventing and controlling obesity.

## Methods

### Literature search strategies and selection

We scoured eight electronic databases for locating research products and policy papers. Initially, we enumerated an exhaustive list of a) research articles by searching on PubMed (US National Library of Medicine) and Google Scholar, and b) reports of nationwide cross-sectional surveys, namely Demographic and Health Survey (DHS), Non-communicable Diseases Risk Factor Survey (STEPS) and Nepal National Micronutrient Status Survey (NNMSS) searching manually on their official websites; published between January 1, 2005 and April 10, 2019. We retrieved obesity-related national policies, strategies and programmes from the websites of Ministry of Health and Population (which implements the government’s health sector nutrition programmes) and the National Nutrition and Food Security Secretariat (which coordinates nutrition activities across half a dozen sectors and maintains a repository of nutrition and food security related policies, strategies and plans). Details of the search terms used and results obtained from these databases are given in Supplementary file 1.

We included studies if: 1) they were published in English between 1 January 2005 and 10 April, 2019; 2) reported prevalence of either obesity or overweight; 3) age of participants was either 0-5 years or 15 years and above. ; and 4) were adequately powered using a sufficiently large sample size. We rendered the studies ineligible if they: 1) did not adopt cross-sectional and population based designs; 2)did not report prevalence of obesity or overweight; 3) they reported prevalence among older children, adolescents or elderly only; 4) were reviews of published articles; 5) analyses of data from secondary sources; or 6) were conducted outside of Nepal (among diaspora Nepali) underpowered studies; and 7) those with any of the key characteristics(design, population, geographical coverage, sampling technique and size) missing were also excluded. However, we included all of the nutrition nutrition-specific policies, strategies and plans promulgated by the federal government in the health or closely related sectors.

### Data extraction and synthesis

The following details were drawn from the included studies: study period (years and months, where available), geographical coverage, study population (gender, age), sampling procedures, sample size, cut-off points used to define overweight and obesity, and prevalences of overweight or obesity (combined), overweight and obesity. Owing to a variation among studies by age of participants, geographical and cultural settings and sampling procedures, we preferred to conduct narrative tabular synthesis over an analysis of individual participant data. Point estimates (prevalence of overweight and obesity) and, where available, interval estimates (95% confidence intervals of the prevalence) were used for such synthesis. Using data from the latest available two reports from surveys conducted on a country-wide scale (DHS and NNMSS), we examined inequalities in overweight and obesity prevalence by gender, geographical locations and household wealth quintiles. Apart from urban and rural areas, we disaggregated prevalences across traditional divisions of eastern, central, western, mid-western and far-western regions; and the new structures of seven provinces. We also calculated urban-to-rural and richest-to-poorest ratios in prevalence of overweight and obesity among adults and children, based on data from the aforementioned two reports.

## Results

### Description of the studies

As illustrated in **Figure 1**, We identified 357 records, which reduced to 336 studies after removal of duplicates. Further, we excluded 259 of them upon screening titles and abstracts. Following a full-text scan for eligibility, we finally incorporated 20 studies (13 peer reviewed articles and 7 survey reports), with a total of 79,082 men and women aged 15 years or more, and 11,866 pre-school children participants.

**Figure 1.**
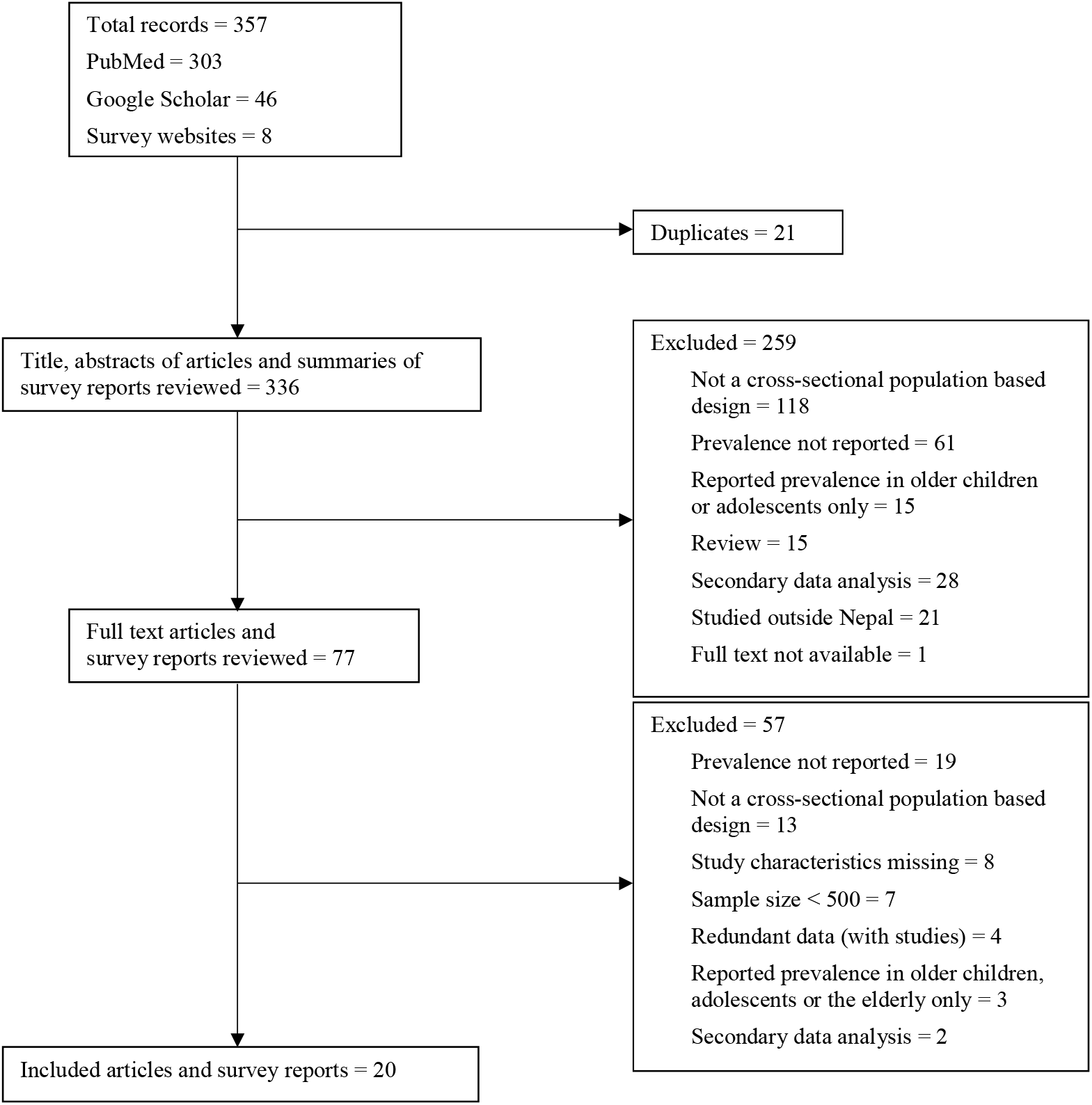
Literature search and selection strategy.

**Table 1** and **Table 2** summarise main features of and findings from the included studies, which are arranged by data collection period (ranging from 2003 to 2017). Age and sex composition of the participants and geographical coverage was not uniform across the studies.. First two of the three rounds of DHSs (2006 and 2011) and NNMSS (2016) incorporated 15-49 years old women and children under five years of age, whereas the latest round of DHSs (2016-17) included both men and women aged 15-49 years and the children.

**Table 1.**
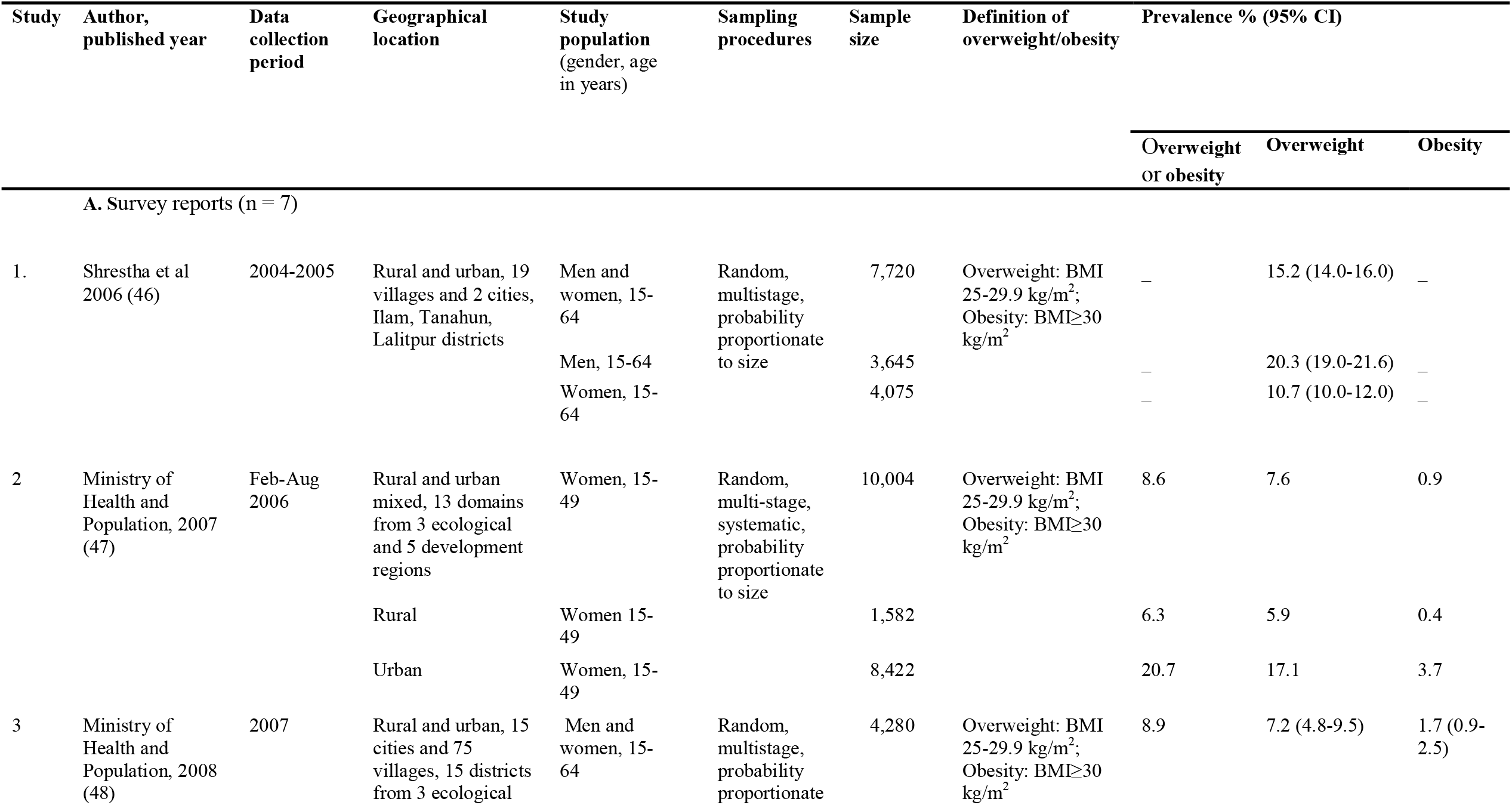
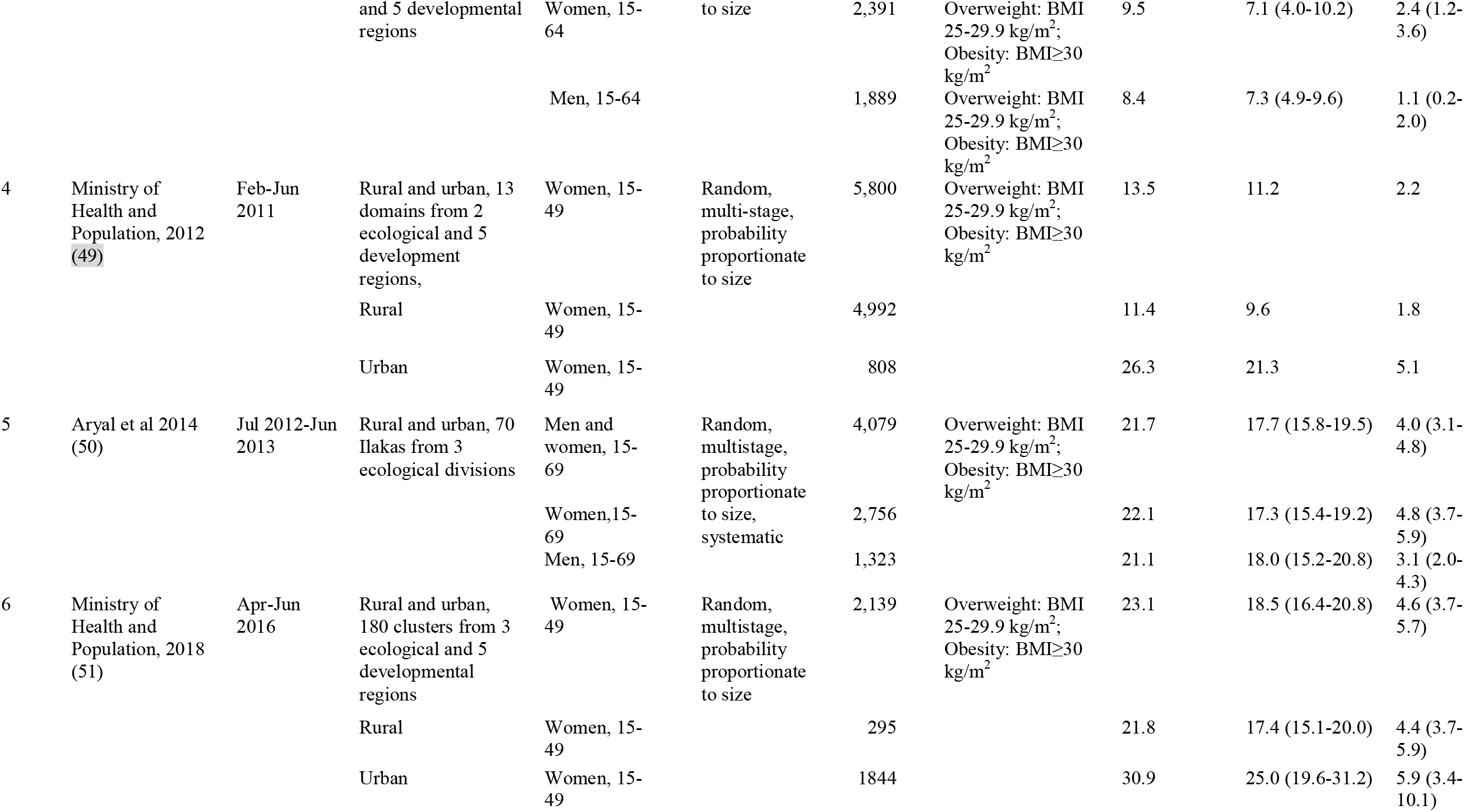
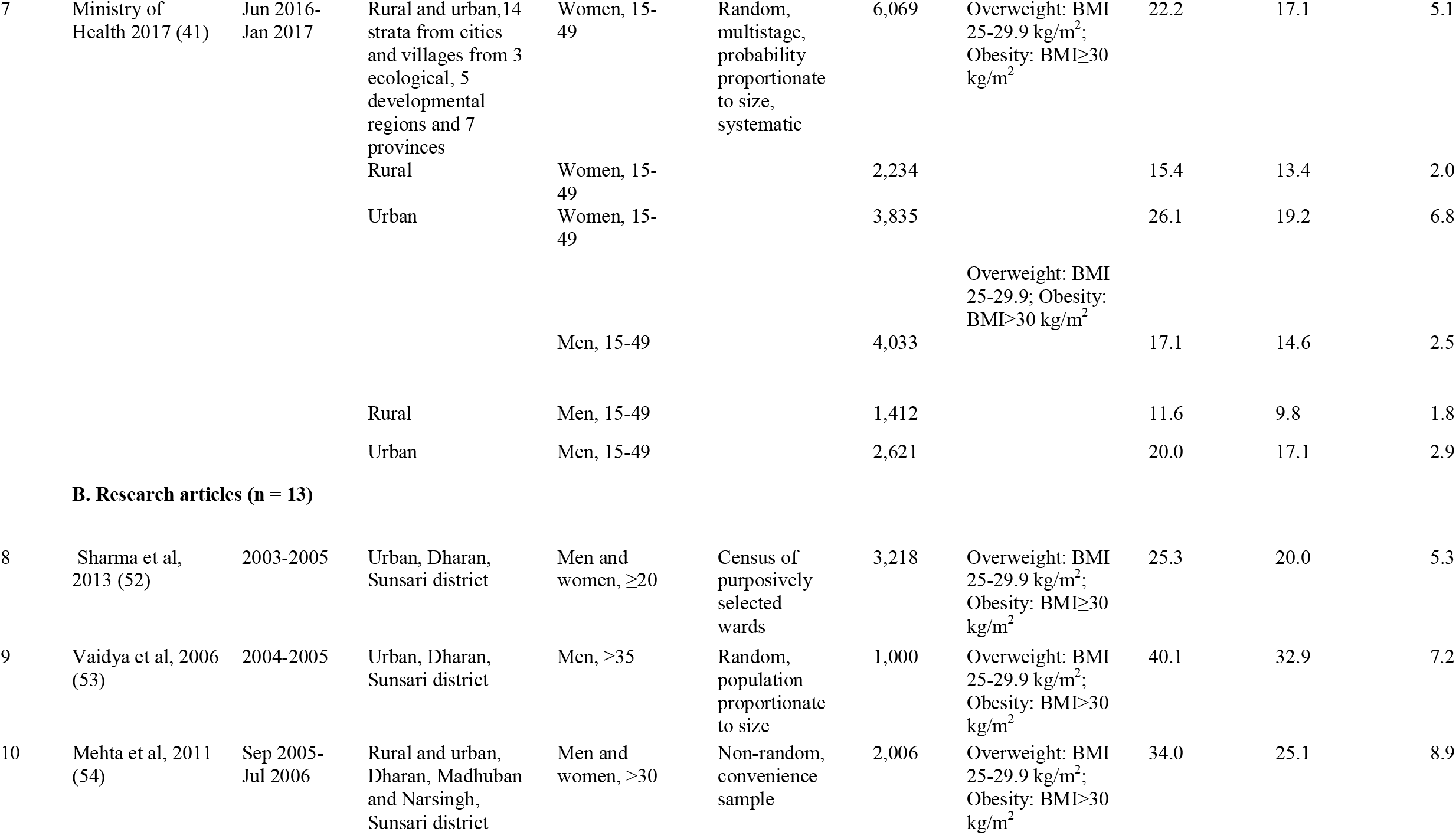
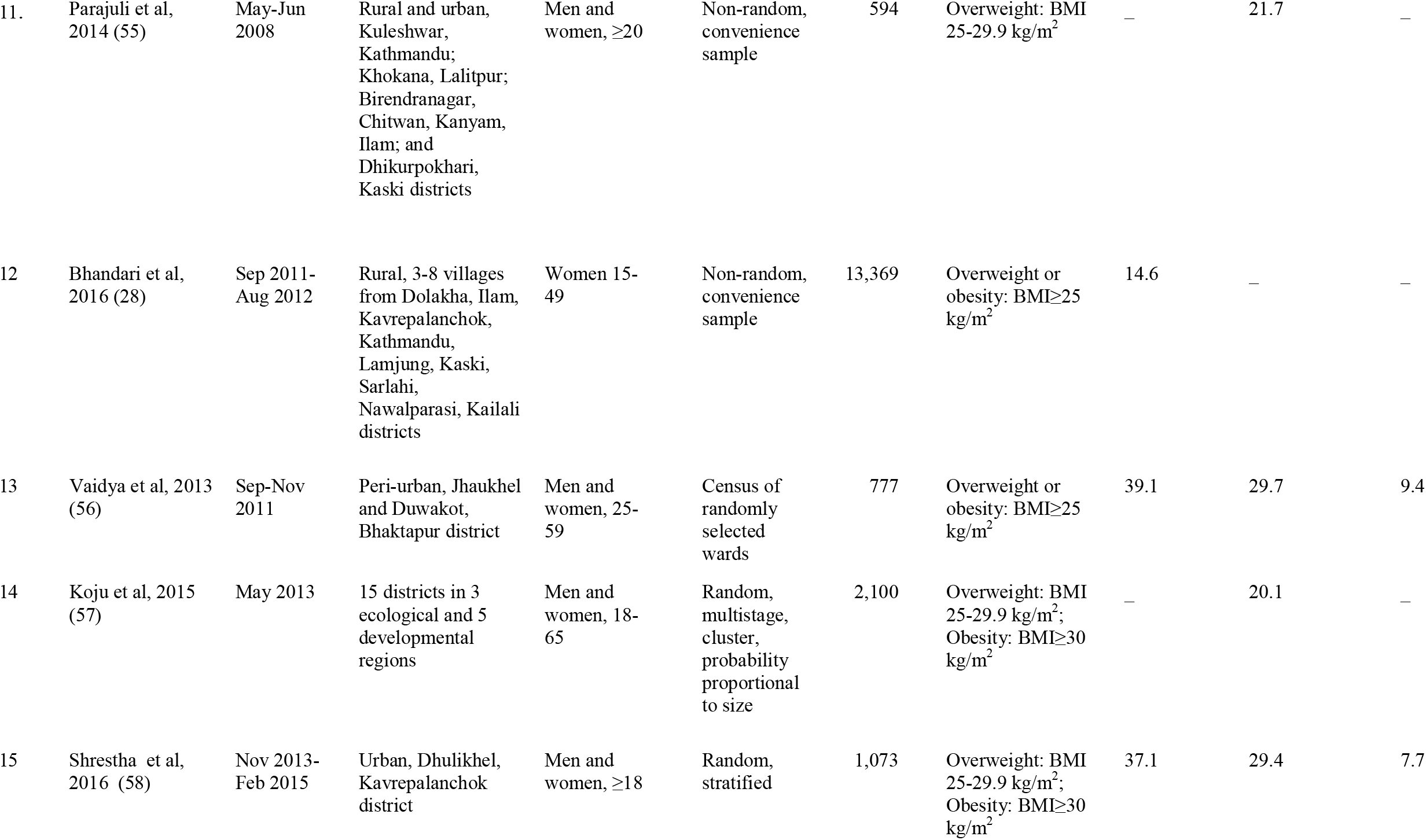
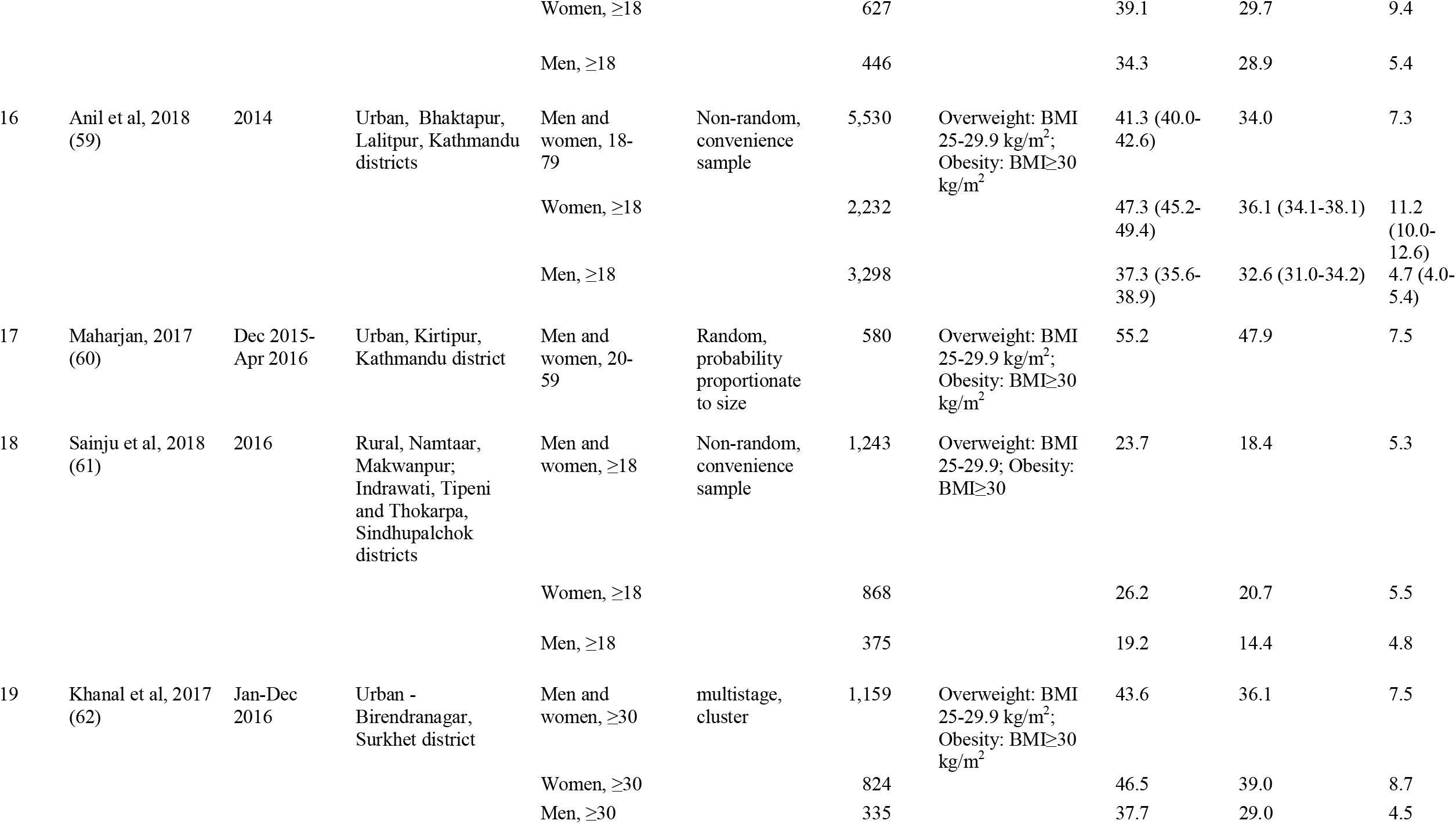
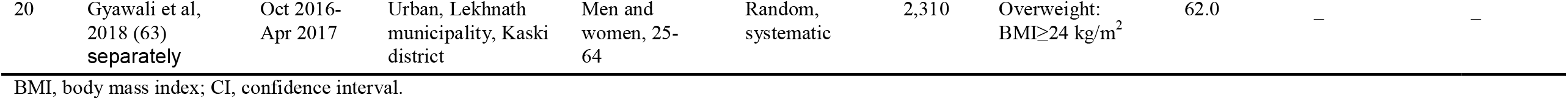
Trends in prevalence of overweight and obesity among people aged 15 years and above in Nepal, 2003-2018.

**Table 2.**
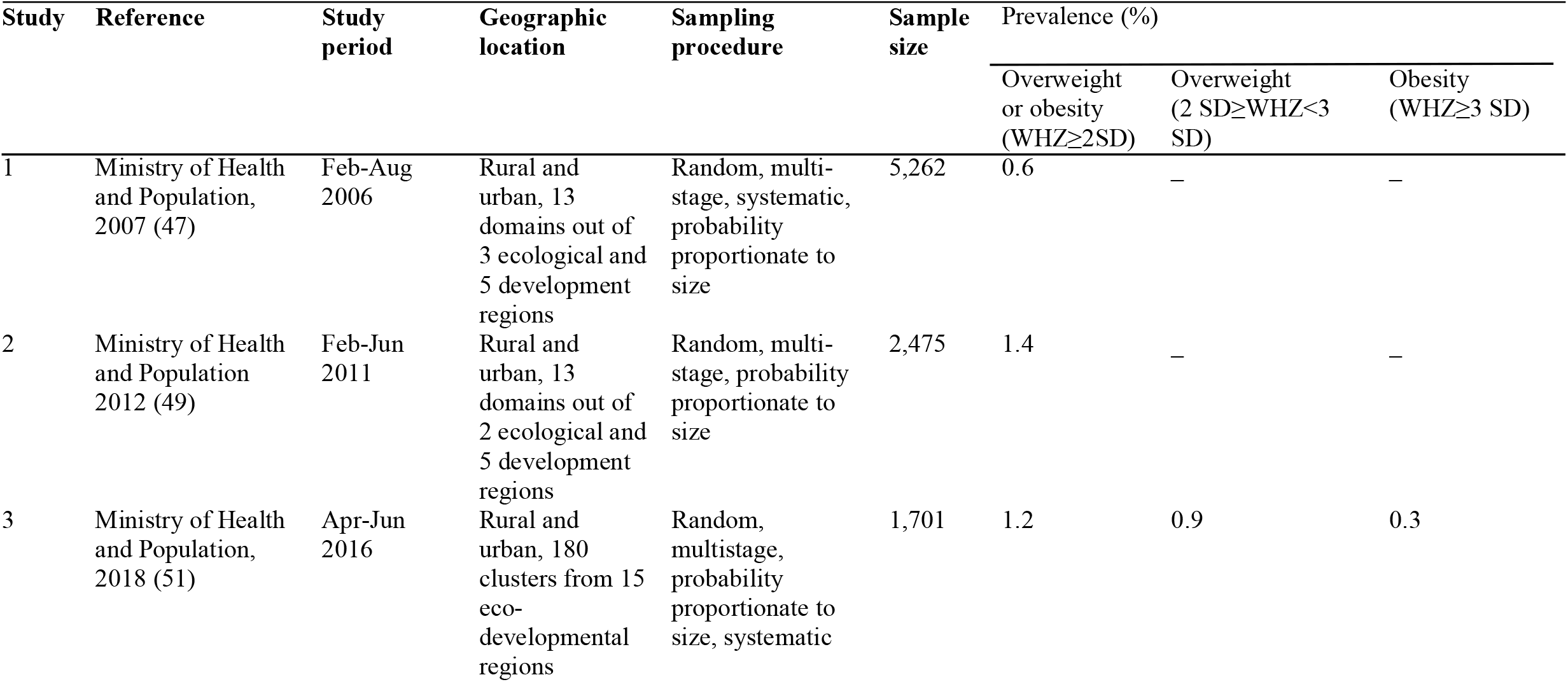
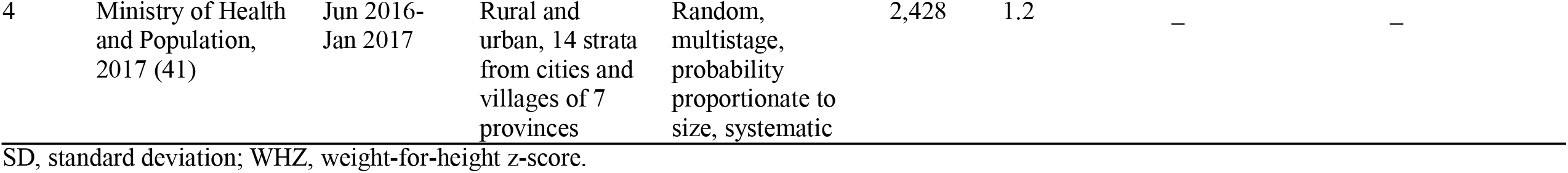
Trends in prevalence of overweight and obesity among children under five years of age in Nepal, 2006-2017.

The STEPS included men and women aged 15-64 years in the first two rounds (2004-05 and 2007) and 15-69 years in the third round (2012-13). Participants in most of the research articles (11 of 13) comprised of men and women with differing age-groups ranging from 18 to 79 years, whereas men aged 35 years or more and women aged 15 to 49 years were included in one each of such research.

Most of the studies enrolled participants from a mix of rural and urban areas (10 of 20) or urban and peri-urban locations only (8 of 20), whereas two of them were conducted exclusively in villages. The DHSs and NNMSS disaggregated prevalence of overweight, obesity and combined overweight or obesity by age, gender, geographical location and household wealth quintiles, while the others reported an aggregate percentage for the entire population or differences by age and gender only. The DHS, STEPS (except for the first round, 2004-05) and NNMSS were conducted on a country-wide scale with samples drawn from all of the hitherto existing five developmental regions (and new divisions of seven provinces in case of the latest round) and the three ecological zones of the country whereas the articles were based on research conducted at varying scales, from all ecological and developmental regions to a single town.

All but one study adopted the World Health Organization’s global standards for classifying overweight and obesity. Overweight and obesity among men and women were defined as 25 kg/m^2^ ≤ BMI < 30 kg/m^2^ and BMI ≥ 30 kg/m^2^, respectively, except one that considered overweight as BMI ≥ 24 kg/m^2^. As for children under five years of age, the Organization’s child growth standards 2006 formed the basis for determining overweight (2 standard deviation (SD) ≤ weight-for-height z-score (WHZ) < 3 SD) and obesity (WHZ ≥ 3 SD).

### Trends in prevalence of overweight and obesity

Overall, the occurrence of overweight as well as obesity increased steadily. Among people aged 15 years or above, the prevalence of overweight rose from 15.2% in 2004-15 to 18.5% in 2016. Obesity alone increased thrice from 1.7% in 2007 to 5.1% in 2016-17. The combined prevalence of overweight or obesity burgeoned more than twice from 8.9% in 2007 to 23.1 in 2016 as illustrated in Table 1. The combined prevalence of overweight or obesity among children remained fairly low, although it doubled from 0.6% in 2006 to 1.2% in 2017 (**Table 2**).

Overweight and obesity were more common among women than men. The latest round of DHS (2016-17) showed that 17.1% of women and 14.6% of men were overweight, respectively as shown in **Table 3**. Similarly, the pace of increment in obesity prevalence among women was much faster (more than five folds between 2006 and 2016) than that among men (slightly over twice between 2007 and 2017). Likewise, the rate of obesity was much higher among women than men (5.1% vs. 2.5%). The 2016 NNMSS reported overweight and obesity rates of 18.5% and 4.6% among women, respectively. (**Table 2**).

**Table 3.**
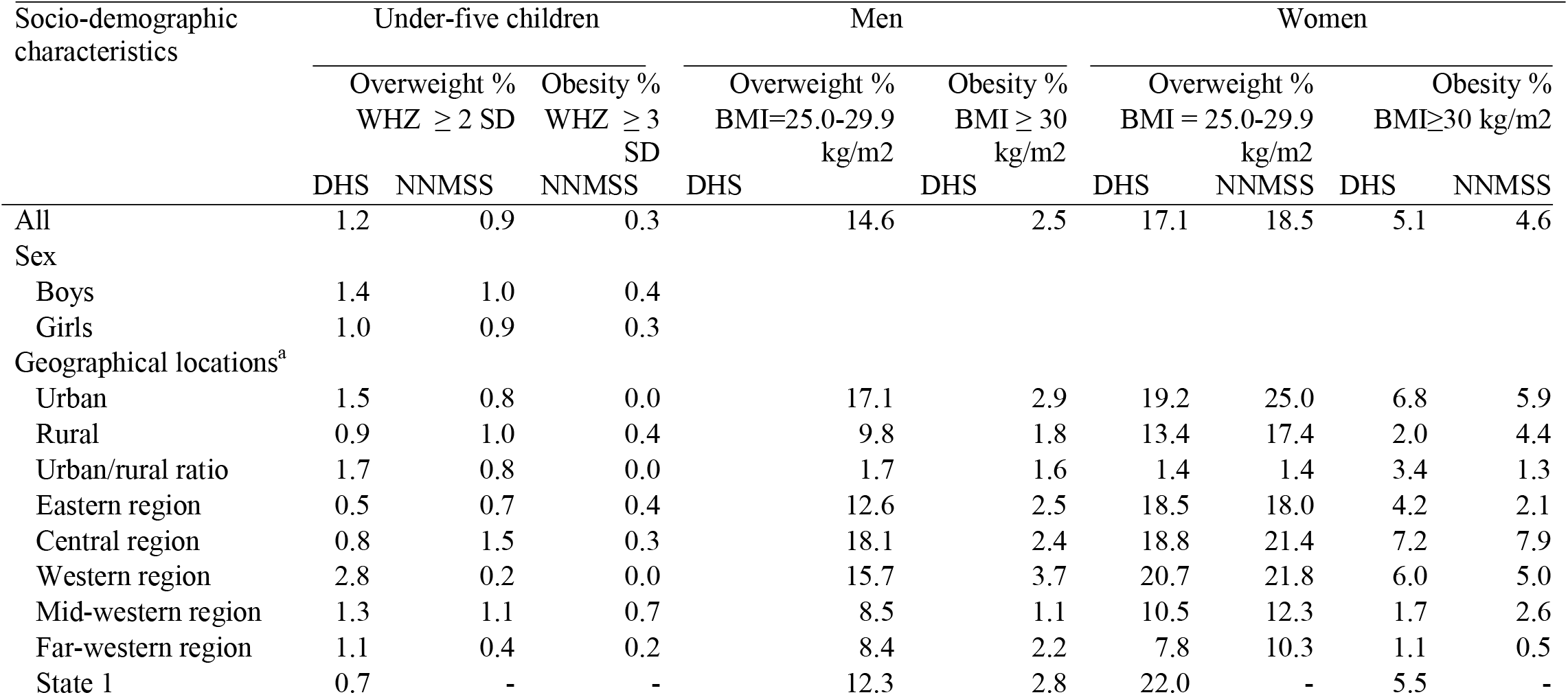
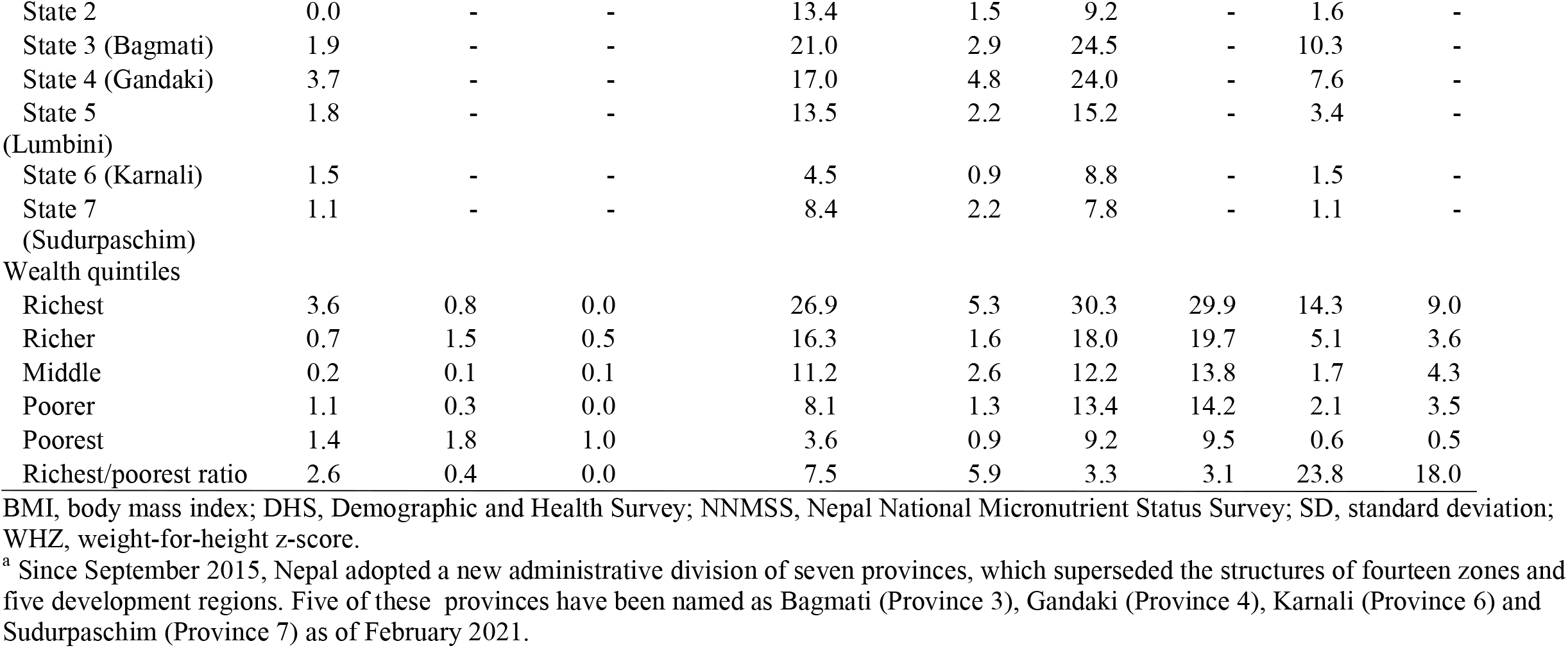
Disparities in prevalence (%) of overweight and obesity across sex, geographic locations and wealth in Nepal based on national survey data from DHS 2016-7 and NNMSS 2016.

| Socio-demographic characteristics | Under-five children |  |  | Men |  | Women |  |  |  |
| --- | --- | --- | --- | --- | --- | --- | --- | --- | --- |
|  | Overweight % |  | Obesity % | Overweight % | Obesity % | Overweight % |  | Obesity % |  |
| | WHZ $\geq 2$ SD | | WHZ $\geq 3$ SD | BMI=25.0-29.9 kg/m2 | BMI $\geq 30$ kg/m2 | BMI = 25.0-29.9 kg/m2 | | BMI$\geq 30$ kg/m2 | |
|  | DHS | NNMSS | NNMSS | DHS | DHS | DHS | NNMSS | DHS | NNMSS |
| All | 1.2 | 0.9 | 0.3 | 14.6 | 2.5 | 17.1 | 18.5 | 5.1 | 4.6 |
| Sex |  |  |  |  |  |  |  |  |  |
| Boys | 1.4 | 1.0 | 0.4 |  |  |  |  |  |  |
| Girls | 1.0 | 0.9 | 0.3 |  |  |  |  |  |  |
| Geographical locations<sup>a</sup> |  |  |  |  |  |  |  |  |  |
| Urban | 1.5 | 0.8 | 0.0 | 17.1 | 2.9 | 19.2 | 25.0 | 6.8 | 5.9 |
| Rural | 0.9 | 1.0 | 0.4 | 9.8 | 1.8 | 13.4 | 17.4 | 2.0 | 4.4 |
| Urban/rural ratio | 1.7 | 0.8 | 0.0 | 1.7 | 1.6 | 1.4 | 1.4 | 3.4 | 1.3 |
| Eastern region | 0.5 | 0.7 | 0.4 | 12.6 | 2.5 | 18.5 | 18.0 | 4.2 | 2.1 |
| Central region | 0.8 | 1.5 | 0.3 | 18.1 | 2.4 | 18.8 | 21.4 | 7.2 | 7.9 |
| Western region | 2.8 | 0.2 | 0.0 | 15.7 | 3.7 | 20.7 | 21.8 | 6.0 | 5.0 |
| Mid-western region | 1.3 | 1.1 | 0.7 | 8.5 | 1.1 | 10.5 | 12.3 | 1.7 | 2.6 |
| Far-western region | 1.1 | 0.4 | 0.2 | 8.4 | 2.2 | 7.8 | 10.3 | 1.1 | 0.5 |
| State 1 | 0.7 | - | - | 12.3 | 2.8 | 22.0 | - | 5.5 | - |
| State 2 | 0.0 | - | - | 13.4 | 1.5 | 9.2 | - | 1.6 | - |
| State 3 (Bagmati) | 1.9 | - | - | 21.0 | 2.9 | 24.5 | - | 10.3 | - |
| State 4 (Gandaki) | 3.7 | - | - | 17.0 | 4.8 | 24.0 | - | 7.6 | - |
| State 5<br>(Lumbini) | 1.8 | - | - | 13.5 | 2.2 | 15.2 | - | 3.4 | - |
| State 6 (Karnali) | 1.5 | - | - | 4.5 | 0.9 | 8.8 | - | 1.5 | - |
| State 7<br>(Sudurpaschim) | 1.1 | - | - | 8.4 | 2.2 | 7.8 | - | 1.1 | - |
| Wealth quintiles |  |  |  |  |  |  |  |  |  |
| Richest | 3.6 | 0.8 | 0.0 | 26.9 | 5.3 | 30.3 | 29.9 | 14.3 | 9.0 |
| Richer | 0.7 | 1.5 | 0.5 | 16.3 | 1.6 | 18.0 | 19.7 | 5.1 | 3.6 |
| Middle | 0.2 | 0.1 | 0.1 | 11.2 | 2.6 | 12.2 | 13.8 | 1.7 | 4.3 |
| Poorer | 1.1 | 0.3 | 0.0 | 8.1 | 1.3 | 13.4 | 14.2 | 2.1 | 3.5 |
| Poorest | 1.4 | 1.8 | 1.0 | 3.6 | 0.9 | 9.2 | 9.5 | 0.6 | 0.5 |
| Richest/poorest ratio | 2.6 | 0.4 | 0.0 | 7.5 | 5.9 | 3.3 | 3.1 | 23.8 | 18.0 |

BMI, body mass index; DHS, Demographic and Health Survey; NNMSS, Nepal National Micronutrient Status Survey; SD, standard deviation; WHZ, weight-for-height z-score.
<sup>a</sup> Since September 2015, Nepal adopted a new administrative division of seven provinces, which superseded the structures of fourteen zones and five development regions. Five of these provinces have been named as Bagmati (Province 3), Gandaki (Province 4), Karnali (Province 6) and Sudurpaschim (Province 7) as of February 2021.

### Inequalities across gender, geographical locations and household wealth

Overweight was twice as common among men as women one and a half decades ago. However, the difference has now reversed as depicted by most recent estimates. Prevalences of both overweight and obesity accelerated faster among women (2.43 and 5.66 times) compared to men (2.0 and 2.27 times) between 2006 and 2016-17, respectively (Table 1).

The latest round of DHS (2016-17) showed that overweight and obesity were more frequently occurring among women (17.1% and 5.1%) than men (14.6% and 2.5%). Such inequality was consistent across rural and urban areas and household wealth quintiles, except for the middle and poorest quintiles where proportion of men having obesity was slighter higher than women. According to the NNMSS (2016) 18.5% and 4.6% of women were having overweight and obesity, respectively.

Although overweight and obesity continue to affect urban dwellers more than the rural residents, the pace of increment of both conditions were faster in the villages compared to the cities and towns. Prevalence of obesity and overweight was much higher in urban than rural areas. For example, 36.1% adults in Birendranagar municipality were having overweight or obesity compared to 18.4% in villages of Makwanpur and Sindhupalchok districts, in 2016. Risk of having obesity among urban women was as much as 3.4 times higher than their rural counterparts.

Latest surveys indicated considerable differences across the developmental regions and provinces. Among the five developmental regions, eastern, central and western regions had much higher prevalence of adult overweight and obesity, relative to mid-western and far-western. Similarly, the rates of overweight and obesity were much pronounced in provinces 1, 3 and 4 than those in provinces 2, 5 and 7 (Figure 2).

**Figure 2.**
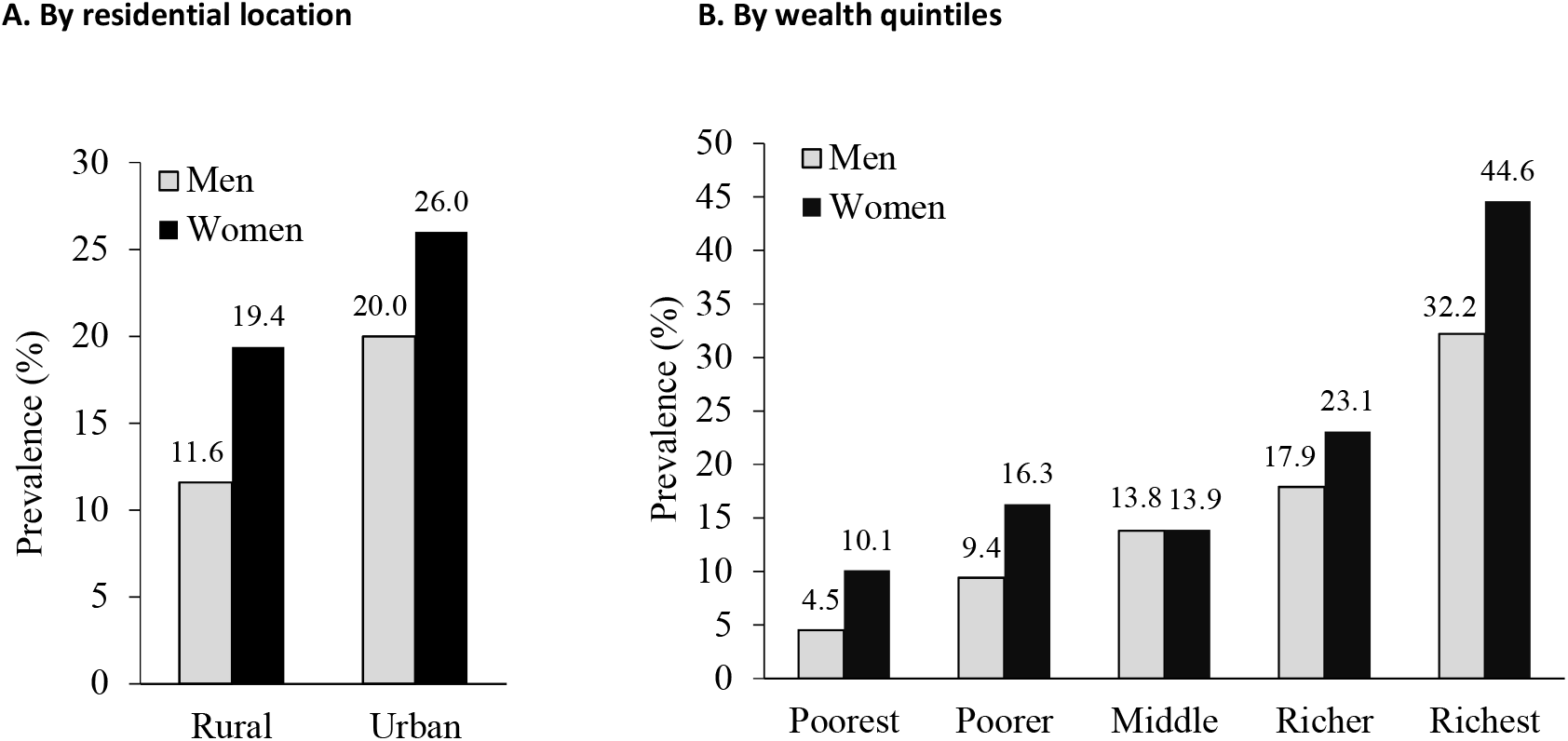
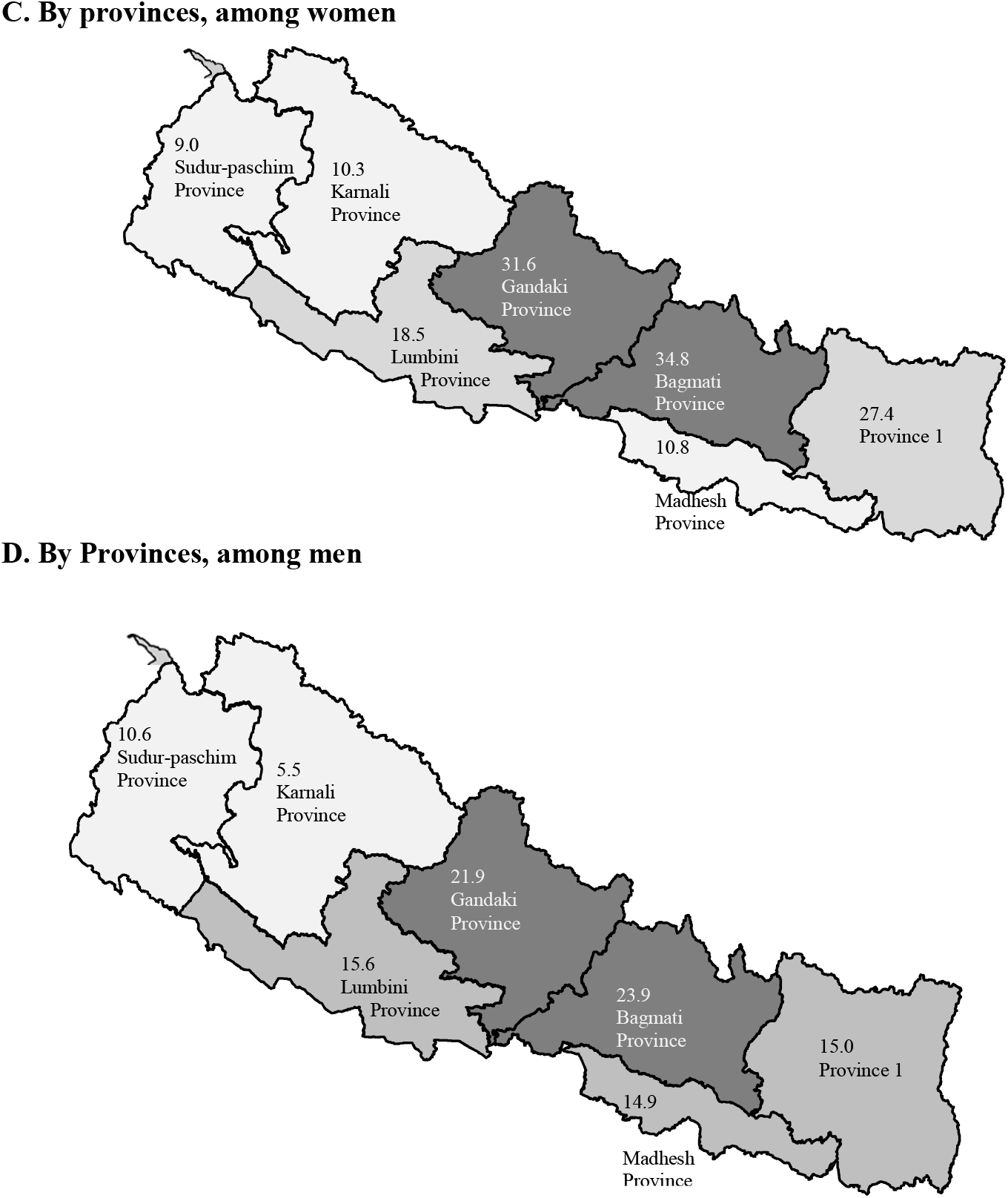
Disparities in prevalence (%) of adult overweight or obesity, based on Nepal Demographic and Health Survey 2016-17.

Overall, the household wealth-obesity/overweight gradient was discernable. The richer had higher rates. Prevalence of overweight among richest men was 7.5 times higher than that among the poorest. Among women, the richest were 18 to 23.8 times more likely to have obesity than the poorest.

In contrast to the situation among adults, the geographical and wealth inequalities in childhood overweight was not consistent across DHS and NNMSS surveys. According to the 2016-17 DHS, prevalence of overweight was greater among boys than girls (1.4% vs 1.0%), 1.7 times higher in urban than in rural areas, and 2.6 times higher among children of the richest than those from the poorest families. However, the 2016 NNMSS did not resonate similar patterns.

### Governmental responses to the obesity epidemic

All of the policies, strategies and plans included in this study were originally designed to mitigate the perennial occurrence of maternal and child under-nutrition, assessed by periodic surveys (such as DHS) in terms of: thinness, anaemia and vitamin-A and iodine deficiency among women; and childhood stunting, underweight, thinness, anaemia, and inadequacy of dietary iodine and vitamin-A. However, they also incorporated some interventions having implications for preventing obesity. We grouped these into direct and indirect preventive or intuitive actions, based on the specific pathways through which they would affect population obesity (**Table 4**). Direct interventions focused on three major dimensions: a) increase the consumption of healthy foods such as fruits and vegetables; b) create environments and promote physical activity; and c) discourage the consumption of sugars, fats and super-processed foods. Indirect interventions included diversification of foods for children, child growth monitoring and nutritional counselling, treatment of child undernutrition, preventing low birth-weight and controlling the use of breast-milk substitutes for infants and young children.

**Table 4.**
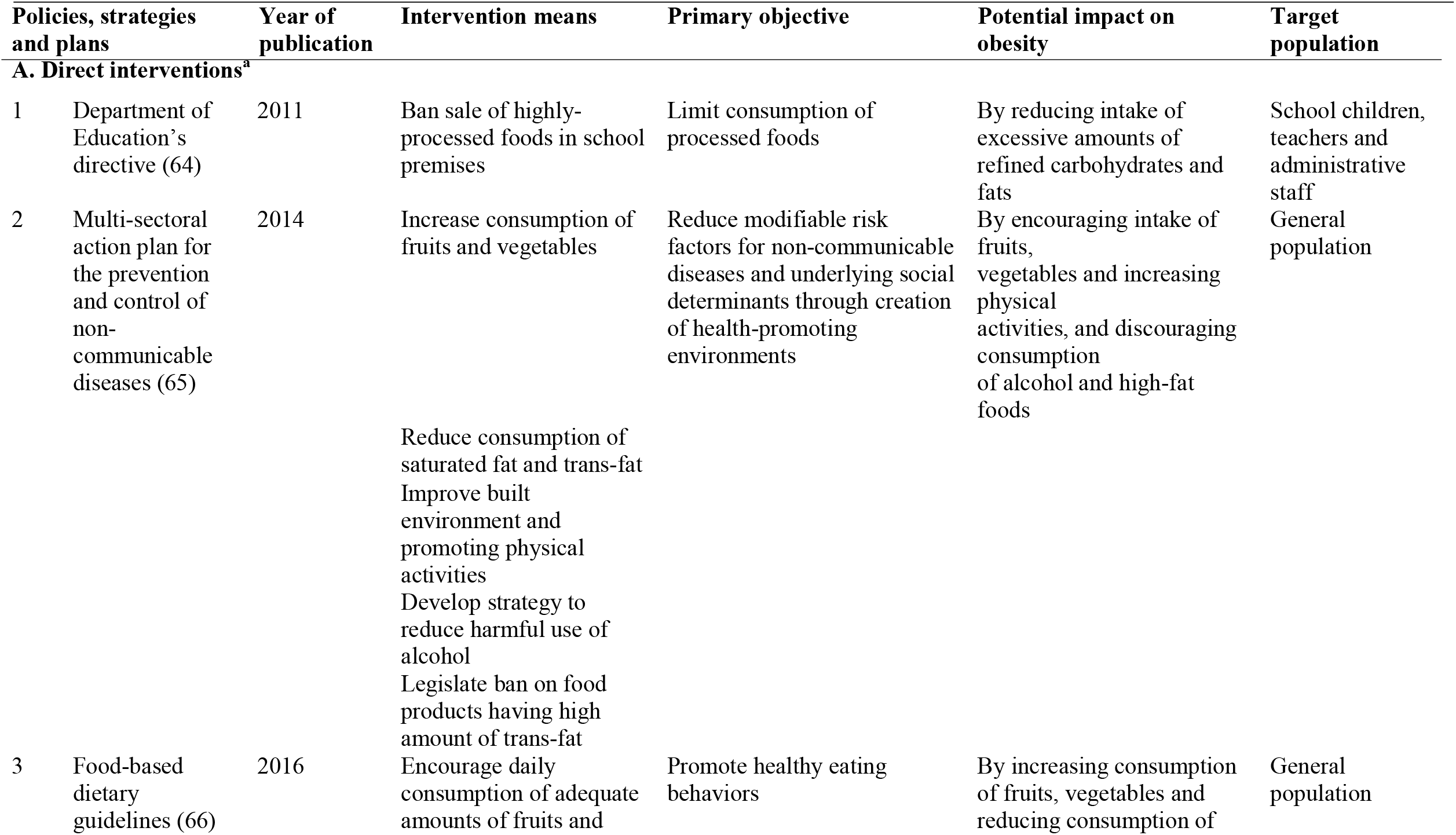
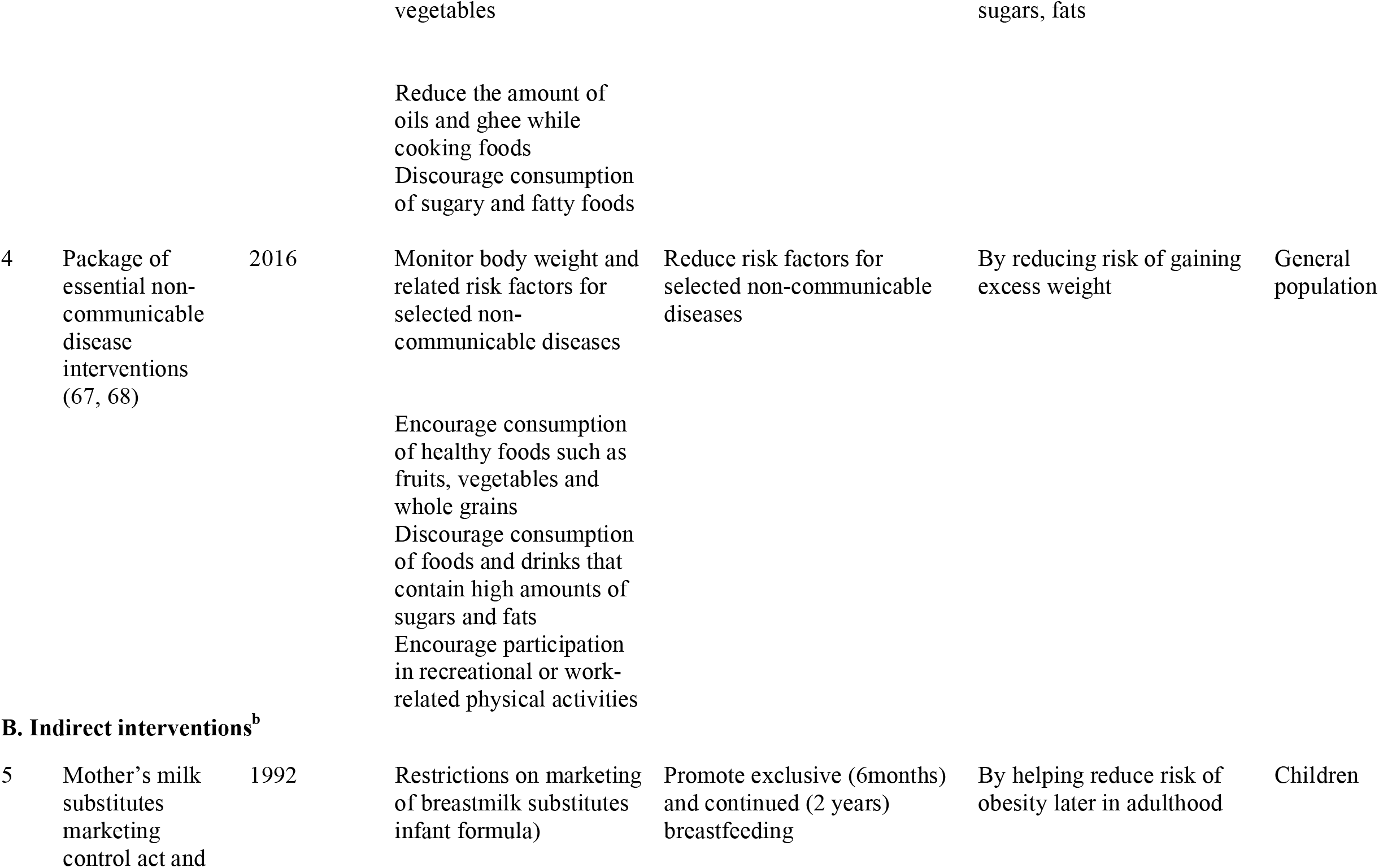
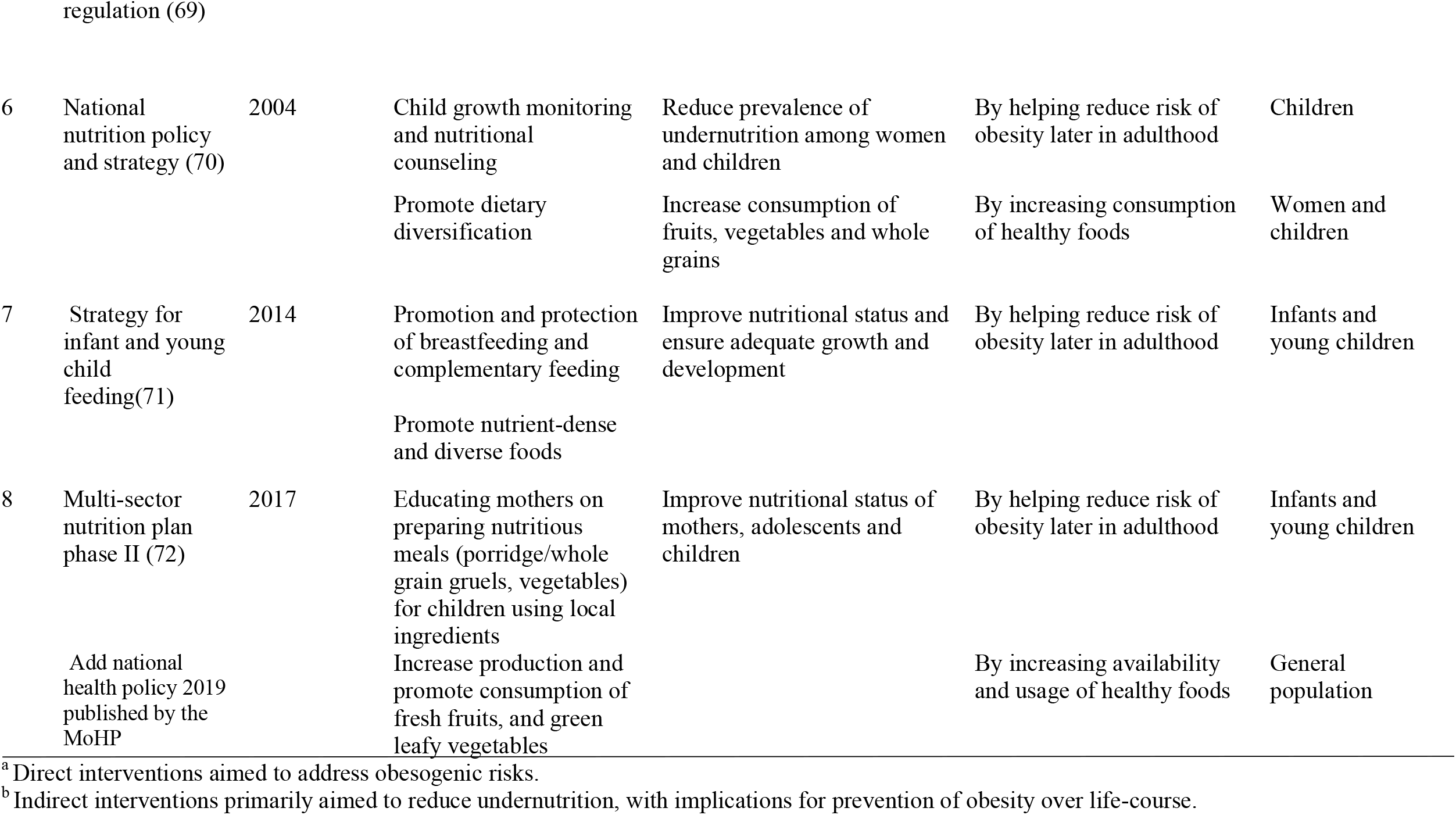
Summary of Nepal government’s nutritional policies, strategies and programmes related to obesity.

## Discussion

Our review illustrates that the prevalence of overweight and obesity among Nepali adults and under-five children had increased substantially across the country since 2004, and hugely in some groups of people. According to latest available data, large disparities existed across groups and regions. Overall, 23.1% of women, 17.1% of men, 1.2% of preschool children had overweight or obesity. In women, the prevalences were 26.0%, 34.8% and 44.6% among urban, residents of Bagmati state and the wealthiest, respectively. Large surveys conducted by the government on a country-wide scale assessed nutritional status (including under-nutrition, overweight and obesity) of children under five years of age and 15-45 year old women only until 2016-17 when men were also included for anthropometric measurements. Data concerning other groups of people such as older children, adolescents, and the elderly are inadequate to infer nationally representative status at the present or trends over the years. Forthcoming research activities should incorporate these groups of the population too, to assist in gauging the transition towards obesity as well as the underlying determinants.

We examined obesity-related national policies, strategies and programs implemented by the government. We identified eight such documents, as listed in Table 4. Much of the focus of existing obesity-related policies, strategies and programmes was to reduce the burden of maternal and child under-nutrition. However, some also aimed directly on promoting healthy eating and physical activity, thus contributing to prevention of obesity. Some others encompassed interventions that have indirect implications for preventing obesity. Yet, the coverage and effectiveness of these interventions have not been evaluated. Research activities in the foreseeable future should endeavor to address this gap.

Rises in the occurrence of population obesity may have resulted from simultaneous changes in people’s income, availability and consumption of foods, and transport. Between 2003-4 and 2015-16, Nepal witnessed a 2.8-fold increment in per capita gross domestic product (32, 33). Annual household expenditure on food rose by 2.4 times between 2003-4 and 2015-16 (34, 35). The number of registered vehicles was 8.5 times more in 2015-16 than that in 2003-04 (36). Changing dietary habits with an increasing availability of highly-processed foods in cities as well as remote villages are also key factors accounting for the propagation of obesity epidemic in this country (37, 38). Similar changes in environmental and socioeconomic factors associated with obesity were observed in many other low- and middle-income countries (16). Our finding of a higher prevalence of obesity among women is also analogous to findings from studies conducted in other South-Asian countries in such as Bangladesh (39).

The higher prevalence of overweight and obesity among urban men and women compared to their rural counterparts is consistent with the situation in most low-income nations, as attested by numerous studies in many countries. However, a highly influential recent analysis of trends based on data from nearly 200 countries concluded that much of the rises in BMI across countries were attributable to increases in villages rather than in cities (40). However, our synthesis of observational studies from Nepal contradicts such finding, thus connoting that the country is in early phases of obesity’s spread into rural and remote regions. Currently higher prevalence of obesity among city-dwelling Nepalis may be attributable to differences with their rural counterparts in terms of several societal and developmental parameters, and associated behaviours. For example, household wealth and food security statuses are higher in urban areas. Moreover, labour-intensive occupations in agricultural production are more common in villages than in the cities where most jobs are physically less demanding (41).

Socio-economic inequalities were underlying the manifest spatial variations in occurrence of obesity. Central and western regions, Bagmati and Gandaki provincess, with higher prevalence of overweight and obesity; were also more urbanised, had lower levels of poverty and achieved better indicators of societal development than the peripheral locations in Lumbini and Sudurpaschim province, for instance (42, 43). This is also corroborated by our results that obesity occurrence was disproportionately affecting wealthier groups of people in all of the states or hitherto existing developmental regions within the country.

Although obesity’s impact on mortality has already surpassed that of undernutrition in Nepal (27), the gamut of governmental nutritional actions is vastly targeted against undernutrition and is inadequate to contain the bourgeoning obesity epidemic. In recent years, several effective strategies have been recommended and utilised elsewhere. These include front-of-pack food labeling; taxing unhealthy foods and subsidising healthier choices such as fresh and wholesome local produce; and improving the quality of food supply, for example by controlling ultra-processed foods and drinks, and increasing availability of natural foods at affordable prices (44, 45). Nepal may need to consider a contextualised implementation of these strategies as integral part of its efforts to control the spread of obesity in the population. Simultaneously, it is also important to enhance the translation existing policies, strategies and plans into practices so as to increase their coverage and effectiveness, for they have incorporated important actions which were either explicitly planned or carry the potential for affecting obesity through fortuitous pathways.

Our findings, however, should be interpreted with caution. We restricted our search databases to PubMed and Google Scholar, which included most of the published research. However, we could not illustrate the full spectrum of Nepal’s obesity situation without the coverage of unpublished studies, which, although difficult to locate, contain results of numerous studies conducted with acceptable levels of precision, given the amount and rigour of prevalence studies conducted by field-based activities of the academic institutions, and a plethora of non-governmental organisations working together with the Nepali state. Second, hhe included reports and articles were based on different sampling procedures, geographical settings and age groups, particularly for adults, thus restricting the calculation of a meaningful aggregate prevalence. Third, all of included studies except one used BMI cut-offs of 25 and 30 rather than the suggested lower limit for certain Asian populations, thereby possibly leading to an underestimation of overweight and obesity prevalences (73). Seven of the eight polices, strategies and plans we selected were promulgated by the government’s health sector (solely, or in partnership with a few other sectors) at the federal level. The programmes and interventions conducted by the health sector at provincial and municipal levels, and other sectors at any level were not adequately covered.

Nonetheless, our study has a couple of comparative advantages over previously published research in this field. It gives an updated description of obesity prevalence over the past one and half decades. The last obesity-specific review from Nepal was published in 2010 (74). In addition, it is the first review of obesity related governmental policies, strategies and plans in the country.

## Conclusion

Put briefly, the prevalence of overweight and obesity increased over the past one and a half decades, particularly, among women, residents of urban and central locations, and the wealthier. Although obesity continues to increase in all sub-groups of the population, large disparities across socio-economic groups and geographical locations continue to exist as illustrated by the latest two country-wide surveys. Governmental interventions were primarily aimed at reducing under-nutrition among women and children, with few actions that directly intervened to prevent the occurrence of obesity. With a persistently high, albeit declining, prevalence of stunting, thinness, underweight and micronutrient deficiencies; governmental efforts to tackle undernourishment may continue to be a priority. Simultaneously, it is important for the state and society to implement effective interventions that would address both the causes and manifestations associated with obesity.

## Data Availability

All data produced in the present study are available upon reasonable request to the authors

## Acknowledgements

We thank Parveen Akhtar (Human Development Research Centre, Pakistan) and Majed Jebril (Ministry of Health, Palestine) for their comments on the literature search strategy and results; Kedar Parajuli (Ministry of Health and Population, Nepal), Anirudra Sharma (Unicef, Nepal), Lonim Prasai (World Health Organization, Nepal) for providing the governmental policies, strategies and plans on nutrition; Peng Nie, Youfa Wang and Junxiang Wei (Xi’an Jiaotong University, China) for criticisms on an earlier plan for and update on the review process; Pawan Acharya (University of Oklahoma) and Naresh Yadav (Ministry of Health and Population, Nepal) for comments on an earlier version of the manuscript.

## Author contributions

Conceptualisation: AB; literature search and selection: AB, DA; drafting and revision: AB, DA. AB and DA have approved the final draft for submission here.

## Notes

### Competing Interest Statement

The authors have declared no competing interest.

### Funding Statement

This study did not receive any funding

### Author Declarations

Data used in this review are publicly available from the the cited articles and the following websites: https://mohp.gov.np http://www.nnfsp.gov.np https://dhsprogram.com https://www.unicef.org/nepal/reports/nepal-national-micronutrient-status-survey-report-2016 https://www.who.int/teams/noncommunicable-diseases/surveillance/data

